# Strengthening Antimicrobial Stewardship in Nigeria: Qualitative and Quantitative Insights into Barriers, Enablers, and Implementation Strategies (2015–2025)

**DOI:** 10.1101/2025.11.04.25339461

**Authors:** Omobolanle Margaret Abe, Rasha Abdelsalam Elshenawy

## Abstract

**Introduction:** Antimicrobial resistance (AMR) poses a major global health threat, particularly in low- and middle-income countries such as Nigeria, where antibiotic misuse is common. Strengthening antimicrobial stewardship (AMS) is essential but remains fragmented and underexplored. This study investigates key barriers, facilitators, and implementation strategies influencing AMS programmes in Nigerian healthcare facilities to inform more effective interventions.

**Methods:** A systematic search of six databases (2015-2025) identified English-language studies on AMS implementation in Nigeria. Eligible studies reported barriers, facilitators, or strategies. Quality was assessed using the Critical Appraisal Skills Programme (CASP) and the Mixed Methods Appraisal Tool (MMAT, 2018). Data on study characteristics, interventions, and outcomes were extracted and synthesised using thematic and quantitative analyses to identify key factors affecting AMS implementation.

**Results:** From 844 screened articles, 10 met inclusion criteria. Major barriers included prescriber resistance (10%), workload (20%), poor infrastructure (40%), policy gaps (50%), limited resources (60%), and inadequate training, with 70% of healthcare professionals showing poor AMS awareness. Facilitators included IT infrastructure (10%), policies (20%), multidisciplinary teams (30%), training (30%), expertise (70%), and leadership support (80%). Common strategies included digital integration, audits, pre-authorisation, education, policy development, surveillance, and AMS committees, outlining a framework to strengthen AMS and reduce AMR in Nigeria.

**Conclusion:** This review highlights substantial challenges to AMS implementation in Nigeria, particularly inadequate diagnostics, limited training, and weak policy enforcement. However, pharmacy-led initiatives and multidisciplinary collaboration show promise. Investment in diagnostic infrastructure, workforce capacity building, policy reform, and sustained leadership commitment is essential to improve AMS effectiveness and combat AMR in Nigerian healthcare systems.

## Introduction

Antimicrobial resistance (AMR) is recognised as one of the greatest threats to global health and development, according to the World Health Organisation [1]. Often referred to as the “Silent Pandemic,” AMR demands urgent action and improved management, and must not be regarded as a problem for the distant future [2, 3]. Global estimates indicate that AMR was directly responsible for more than 1.2 million deaths in 2019, with projections suggesting this could rise to nearly 10 million deaths annually by 2050 without decisive action [4, 5]. According to the UK’s O’Neill Report (2016), antimicrobial resistance (AMR) could surpass all other causes of death worldwide by 2050 if effective preventative measures are not implemented. Nigeria faces a critical situation with AMR [6]. Already burdened by inadequate water, sanitation, and infection control, Nigeria is seeing a rise in resistant infections [7]. Antibiotic use among hospitalised patients in Nigeria is estimated at 65%-79% which is alarming, especially given the limited development of formal antimicrobial stewardship programmes [8]. AMR is not just a human health issue in Nigeria, it is spreading through animal populations and the environment, underscoring the urgent need for a unified One Health approach that integrates human, animal and environmental health strategies [9].

Antimicrobial Stewardship (AMS) is an important public health strategy that promotes the responsible use of antimicrobial medications to enhance patient outcomes, reduce AMR, and maintain the efficacy of existing treatments [10]. AMS uses strategies like choosing the right drugs, doses and treatment lengths to fight infections effectively while reducing side effects and resistance following the WHO’s AWaRe framework [11]. However, Nigeria has struggled with AMS implementation. A 2018 survey showed that only 35% had a formal AMS team and 12% had audited antibiotic use or reviewed prescriptions after 48 hours [12]. This is made worse by poor prescribing habits, easy access to antibiotics without prescriptions, weak regulations and lack of awareness among healthcare workers and the public [13]. Regulatory bodies like NAFDAC and the Pharmacy Council of Nigeria (PCN) are responsible for quality control and professional standards [14]. Therefore their ability to stop misuse and counterfeit drugs remains inconsistent.

To address these gaps, Nigeria created the National Action Plan on AMR (2017– 2022), led by the Nigeria Centre for Disease Control (NCDC). A new version, AMR 2.0, launched in 2024, brings Nigeria in line with global efforts against antimicrobial resistance, including the United Nations (UN) declaration [15]. Key stakeholders in this fight include the healthcare professionals (doctors, pharmacists, lab staff) who prescribe, dispense, and diagnose; policymakers and regulators like NCDC, NAFDAC, and PCN; healthcare facilities at all levels (tertiary, secondary, and primary); community pharmacists and patent medicine vendors; and the public and environmental health sectors, which address the role of animals and the environment in AMR [16].

Although Nigeria has national policies to address AMR, putting them into practice is still a challenge. Studies show that many doctors and healthcare workers do not know enough about antibiotic stewardship, and there are problems with monitoring, training, and following guidelines in healthcare facilities [17]. To improve this, it is essential to thoroughly examine what is stopping or helping the use of AMS in Nigerian healthcare. Current information is incomplete and varies from place to place. This review will examine the implementation of AMS in Nigeria, with the aim of providing policy recommendations and guidance to strengthen future implementation efforts.

## Materials and Methods

### Registration

The systematic review was registered with PROSPERO (registration number CRD420251165641) prior to the commencement of the search process. The PICO (Populations, Intervention, Comparison and Outcome) framework guided the search strategy as shown in Table 1. A comprehensive search of relevant databases was conducted using a combination of relevant keywords and synonymous terms (further details provided in Table 2 and 3). The review followed the Preferred Reporting Items for Systematic Reviews and Meta-Analyses (PRISMA) 2020 guidelines to maintain methodological precision. This PRISMA 2020 served as a guidance to systematically review relevant studies published between 2015 and 2025. Since this study analysed existing published data, formal ethics approval wasn’t needed. However, standard ethical practices for systematic reviews were followed, including proper citation, avoiding plagiarism, and respecting intellectual property.

**Table 1:**
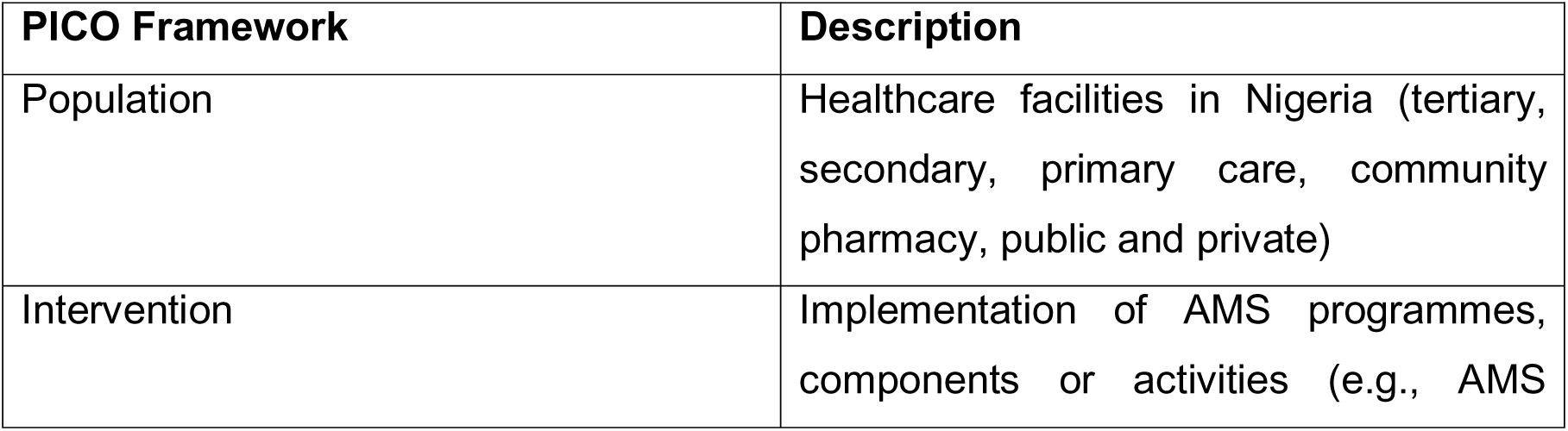
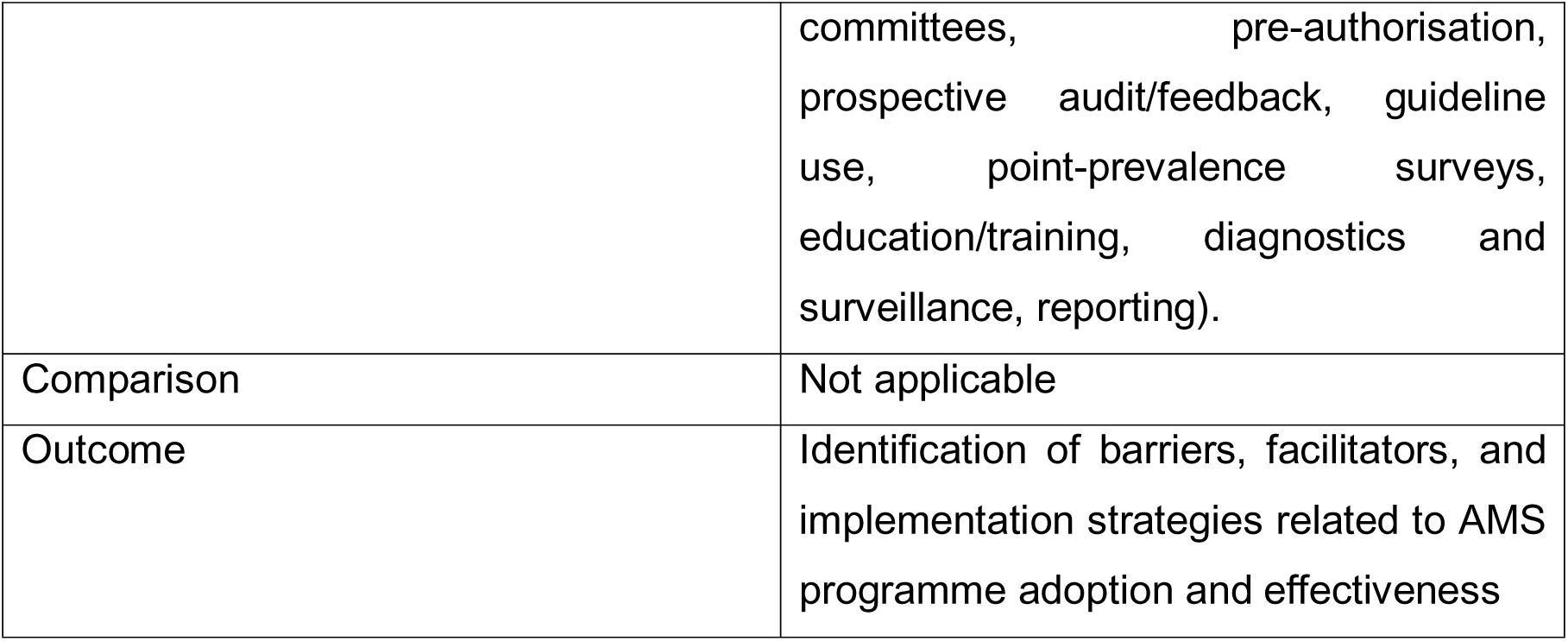
PICO framework and description.

| PICO Framework | Description |
| --- | --- |
| Population | Healthcare facilities in Nigeria (tertiary, secondary, primary care, community pharmacy, public and private) |
| Intervention | Implementation of AMS programmes, components or activities (e.g., AMS |
|  | committees, pre-authorisation, prospective audit/feedback, guideline use, point-prevalence surveys, education/training, diagnostics and surveillance, reporting). |
| Comparison | Not applicable |
| Outcome | Identification of barriers, facilitators, and implementation strategies related to AMS programme adoption and effectiveness |

**Table 2:**
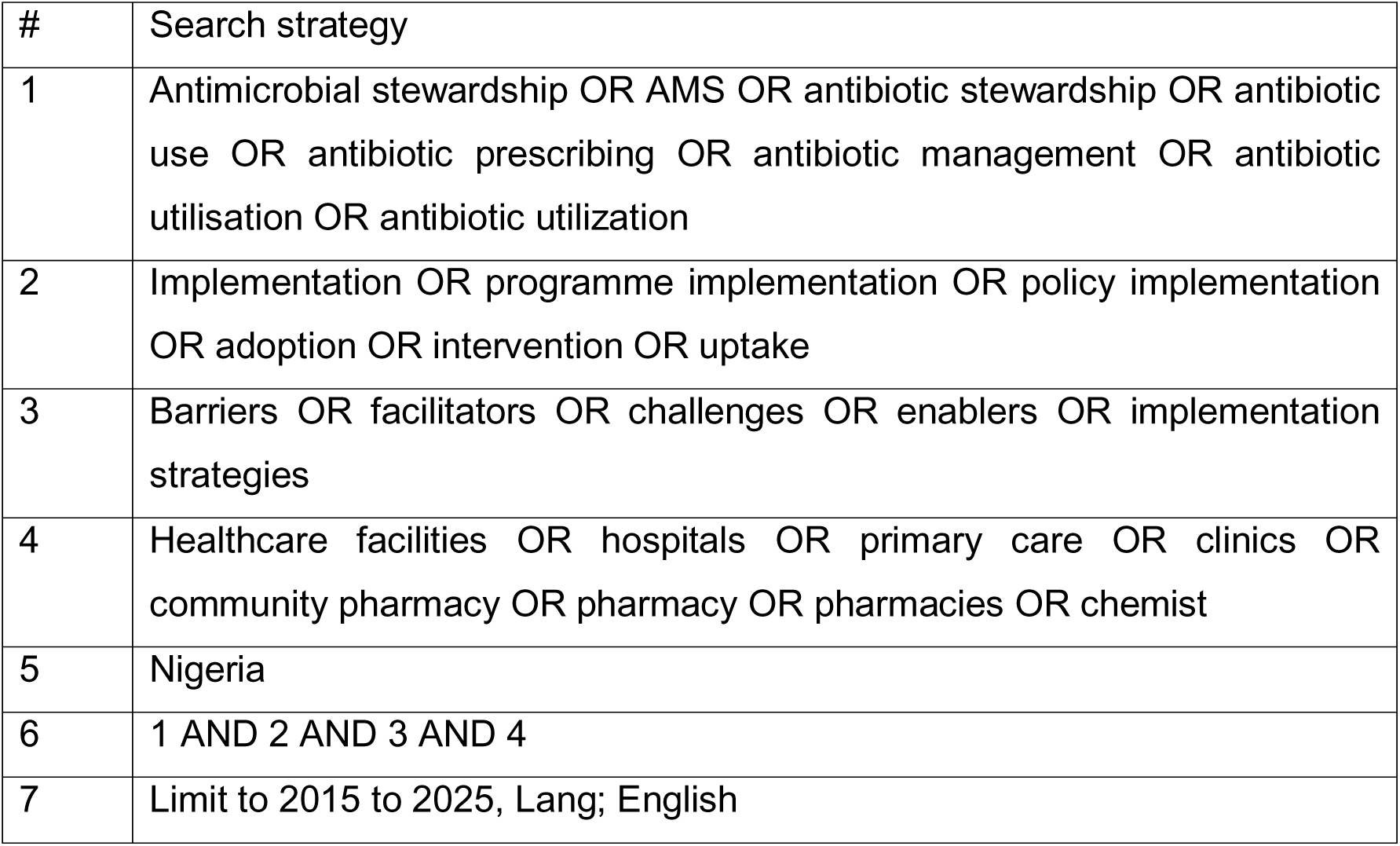
Systematic literature review of search strategies.

**Table 3:**
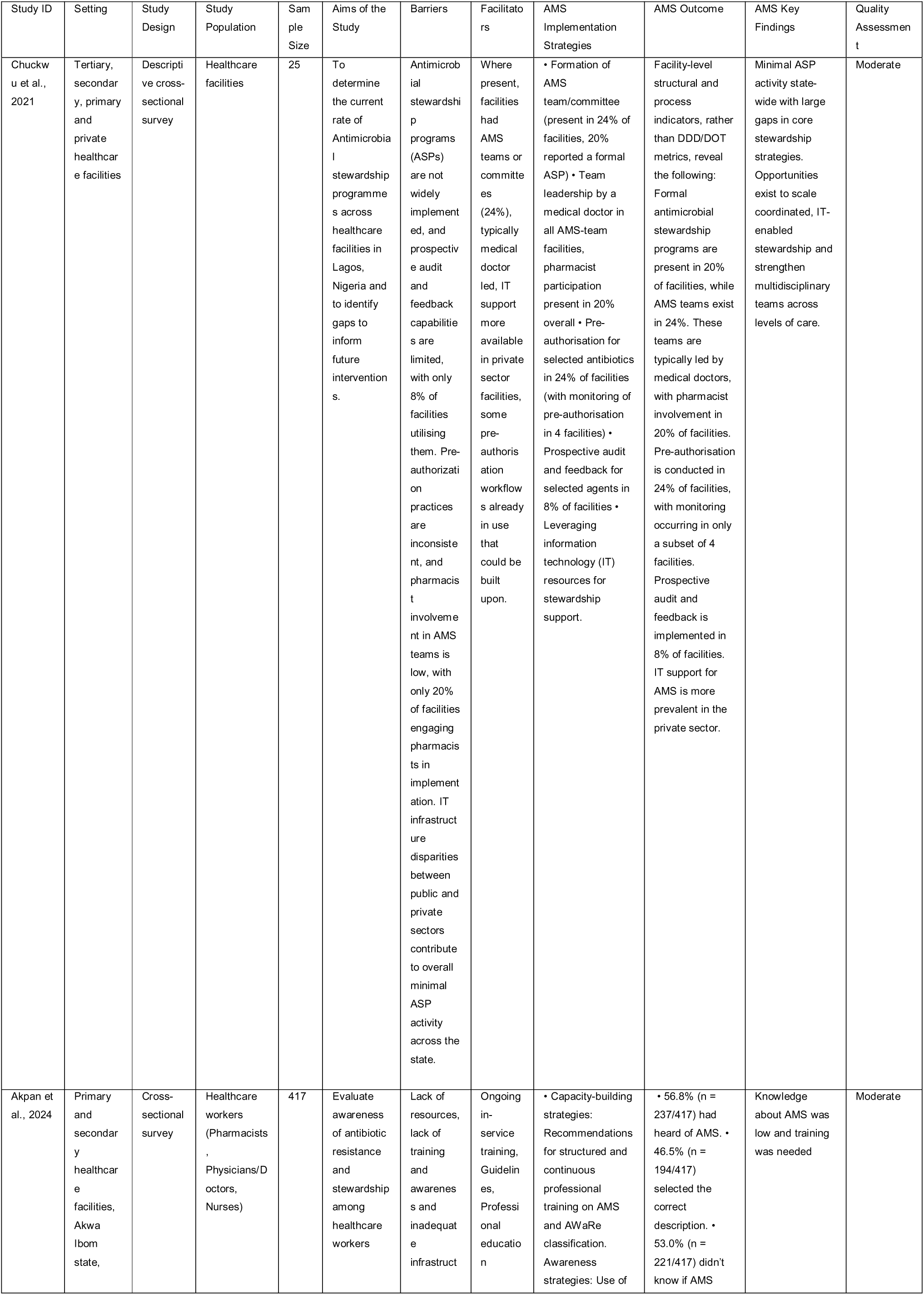
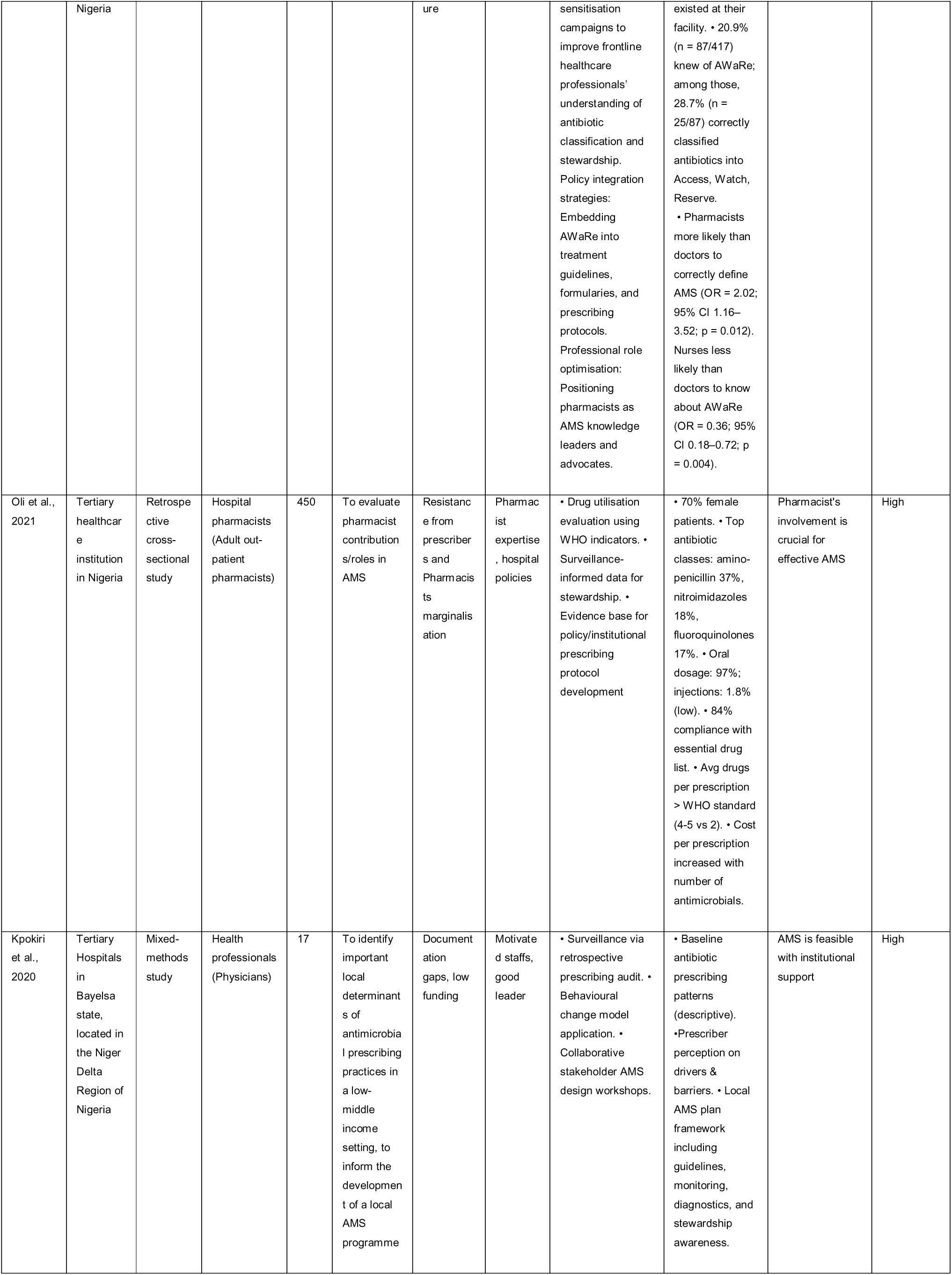
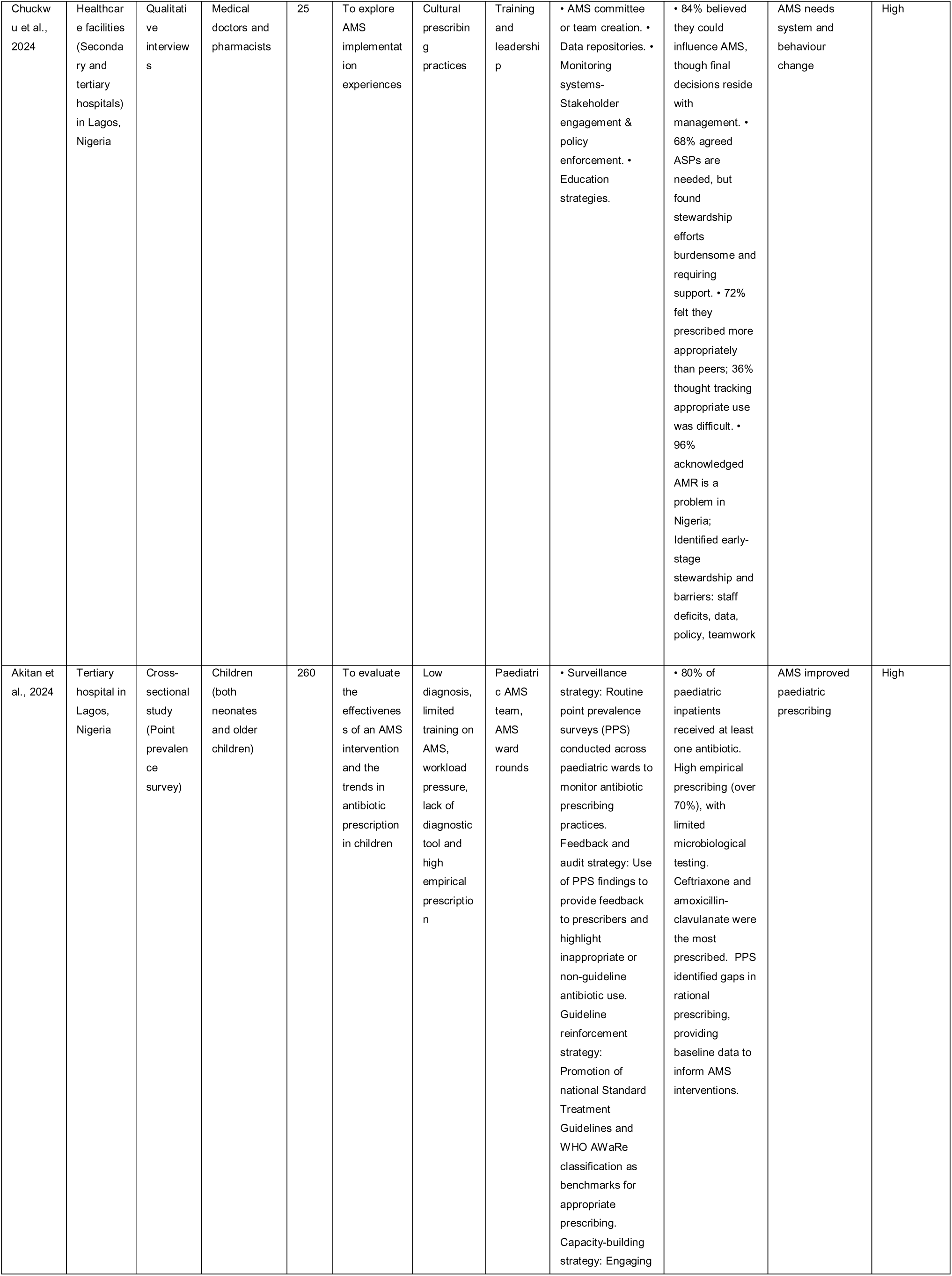
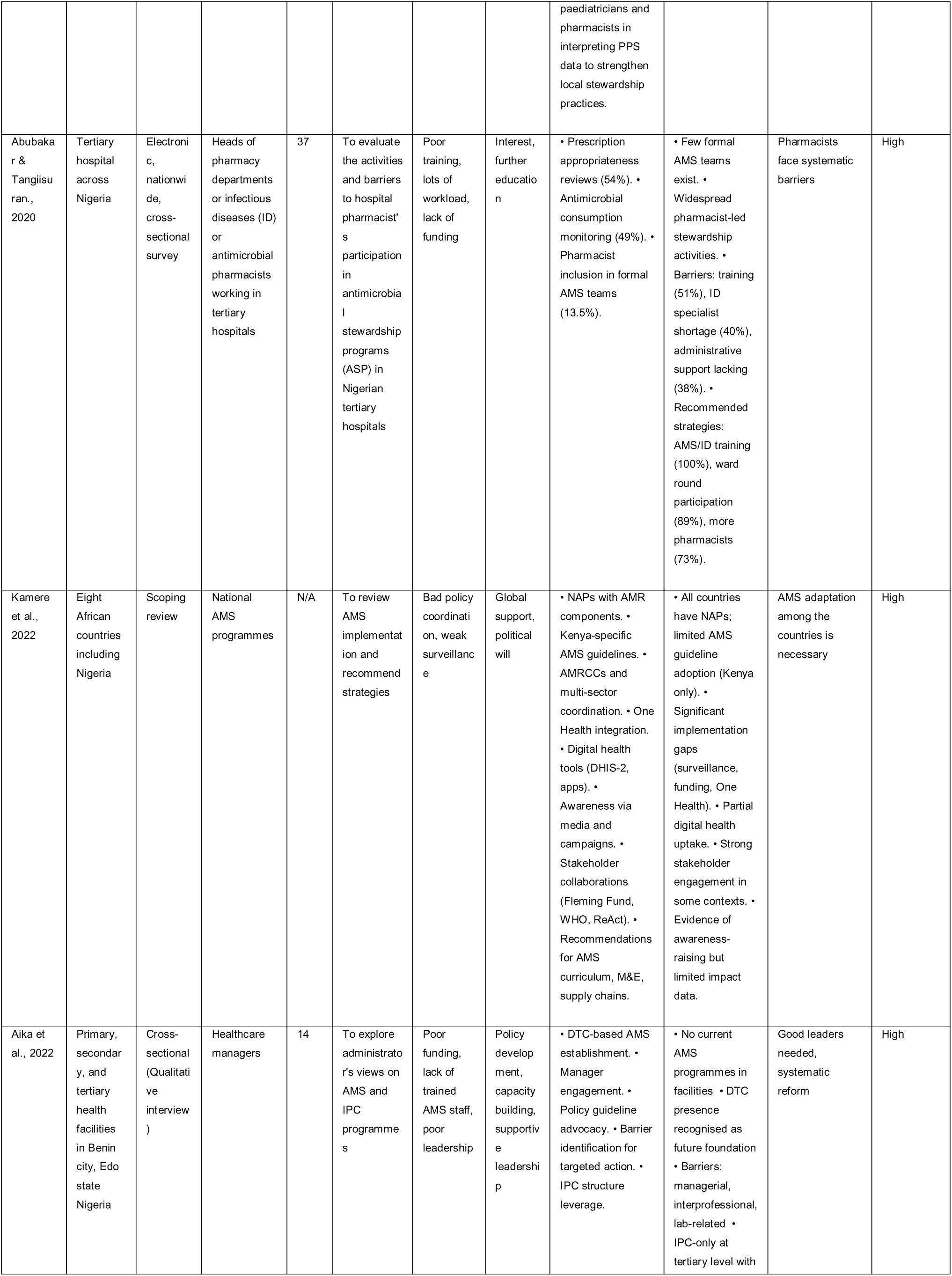
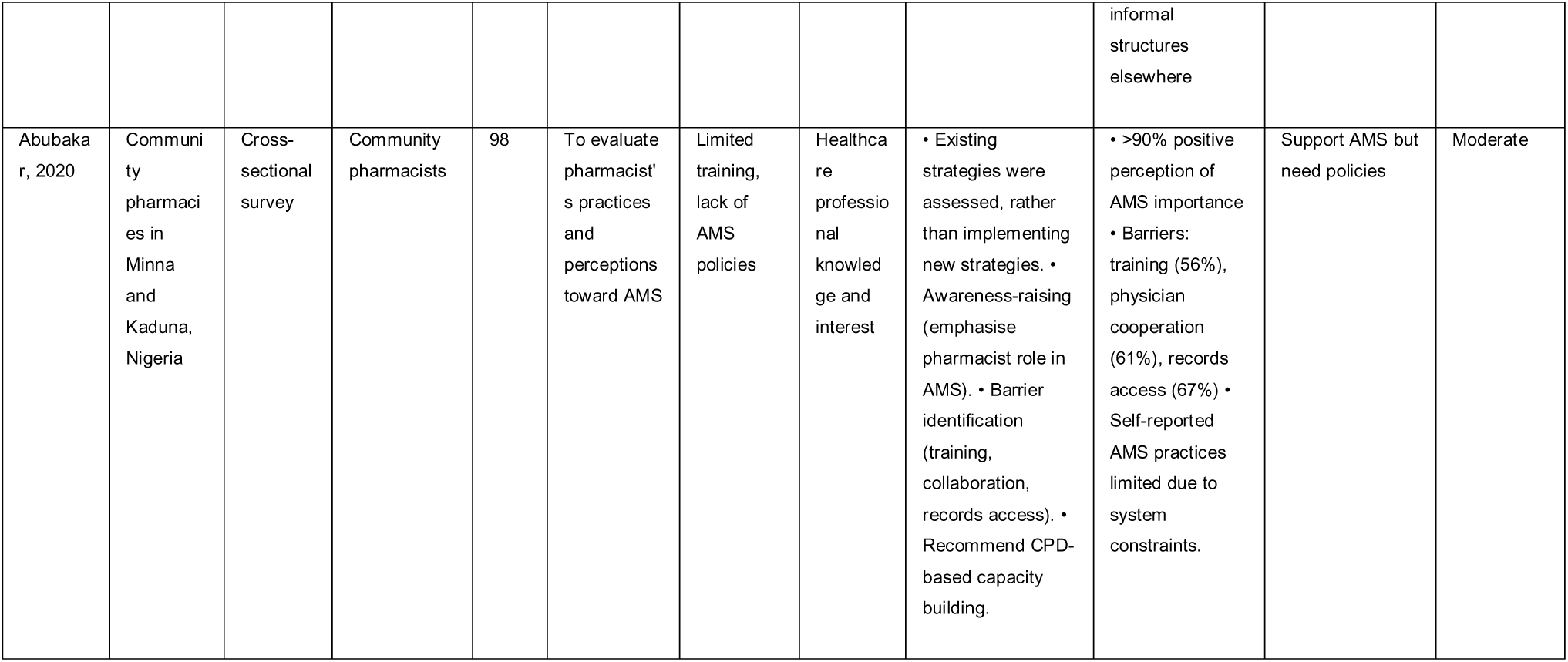
Data extraction table for included studies.

### Eligibility Screening

The articles retrieved from the databases were exported to Excel sheets for screening and identification of the eligible articles. Titles and abstracts were screened for relevance; duplicates were removed, followed by a screening of the complete articles for possible inclusion.

#### Inclusion Criteria

Selected studies were assessed against the following inclusion criteria: (i) Studies published in English (ii) Conducted in Nigeria, in any healthcare facility (tertiary, secondary, primary care; public or private) (iii) Report on the implementation, evaluation, or description of antimicrobial stewardship (AMS) programmes, interventions, or activities (iv) Peer-reviewed qualitative, quantitative, or mixed-methods studies (v) Implementation research, programme evaluations, or observational studies (vi) Published between 2015 and 2025 (vii) Report barriers, facilitators, or strategies related to AMS implementation

#### Exclusion Criteria

(i) Studies not focused on healthcare facility settings (e.g., purely community based studies without AMS components) (ii) Studies not addressing AMS implementation (e.g., those focused only on resistance surveillance, prescribing patterns, or infection control without AMS relevance) (iii) Commentaries, editorials, conference abstracts, letters, or opinion pieces without original empirical data (iv) Non-English publications (v) Studies conducted outside Nigeria

#### Data sources and search method

A systematic search was conducted using PubMed, Scopus, Google Scholar, Cochrane Library, CINAHL, and the Web of Science. Using Boolean operators (Table 2), keywords and MeSH terms like “antimicrobial stewardship,” “implementation,” “barriers,” “facilitators,” “healthcare facilities,” “Nigeria,” and “community pharmacy” were added. The search was limited to English language studies published between 2015 and 2025.

### Quality Assessment of included studies

The quality of the studies included was assessed using the Critical Appraisal Skills Programme (CASP) checklists and the Mixed Methods Appraisal Tool (MMAT, 2018 version). CASP was used for qualitative and observational studies, focusing on aims, methods, data, ethics, analysis, and clarity. MMAT was applied to mixed-methods studies to evaluate qualitative, quantitative, and integrative elements. This combined approach ensured rigour despite study differences. While CASP and MMAT improved assessment reliability, some subjectivity in interpreting criteria remained.

### Data extraction and analysis/synthesis

#### Data Extraction

A detailed form was created in Microsoft Excel to extract key study information, such as; author, year, study design, study setting, study population, sample size, aims/objectives of study, barriers to AMS implementation, facilitators of AMS implementation, AMS implementation strategies, AMS outcomes or metrics and measures, AMS key findings, and quality appraisal using CASP & MMAT tools (Table 3). Data extraction was performed independently and verified to minimise bias. Data obtained were summarised using narrative synthesis and thematic analysis.

#### Data Analysis and Synthesis

Due to the variety of study designs, a narrative synthesis and thematic analysis were used for qualitative data. This allowed for the integration of different types of evidence while maintaining detailed analysis. Barriers and facilitators were coded into themes like infrastructural, human resource, behavioural, and policy-related factors. Strategies were mapped against AMS frameworks such as the WHO core elements for hospital AMS programs. This method effectively captures detailed contextual insights, but it may have some subjectivity in coding.

## Results

The search processes identified a total of 844 results from various databases including PubMed (n = 196), Scopus (n = 370), Web of Science (n = 48), Cochrane Library (n = 25), CINAHL (n = 7) and Google Scholar (n = 198). After removing duplicates, 694 articles were left for title and abstract screening. Out of these, 76 published articles were eligible for full-text review, with 56 meeting the inclusion criteria (Figure 1). Forty-six articles were excluded for not meeting the criteria due to reasons such as the study not focusing on AMS implementation (n = 14), not in Nigeria (n = 21), or not in a healthcare setting (n = 11). Ultimately, 10 studies were included in the final analysis (Figure 1).

**Figure 1:**
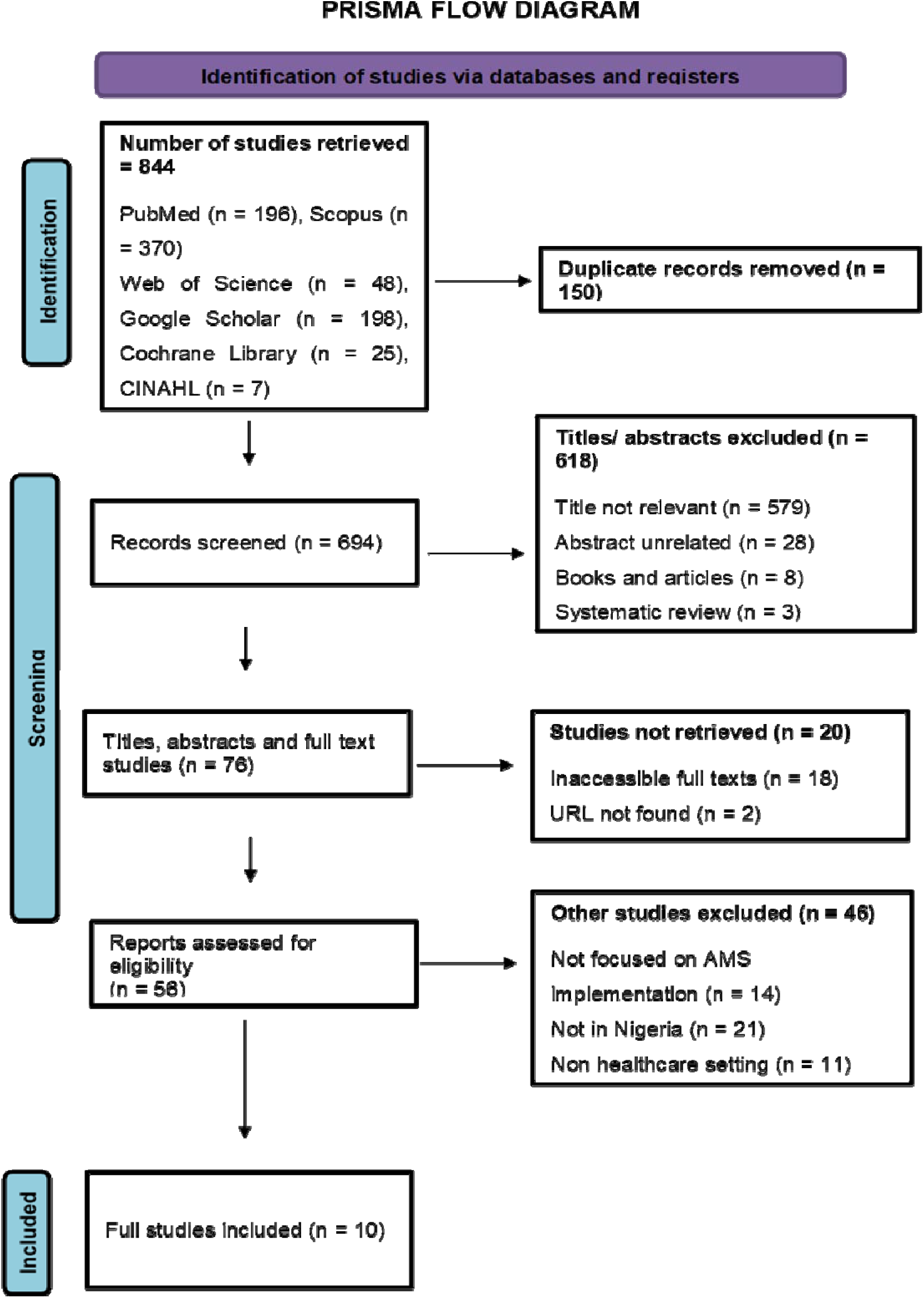
PRISMA flowchart of eligible studies for inclusion in the systematic review. (Retrieved 05/08/2025)

### Quantitative analysis

#### Characteristics of Included Studies

A total of 10 studies, conducted between 2015 and 2025 met the inclusion criteria, encompassing 1,343 participants across Nigerian healthcare facilities. The studies demonstrated a broad geographical representation, spanning all six geopolitical zones, with Lagos State contributing the highest proportion (30%) and several studies adopting nationwide coverage. The majority of studies were set in tertiary hospitals (40%) and mixed-level facilities (40%), while only a limited number explicitly involved primary healthcare settings. In terms of methodological designs, cross-sectional surveys were the most common (40%), followed by qualitative interview studies (20%), with the remainder using mixed or descriptive approaches (Table 3). Study participants were primarily healthcare professionals, with pharmacists featuring in 60% of studies, highlighting their central role in AMS implementation. Sample sizes varied widely, ranging from small qualitative studies (≤25 participants) to large surveys (>300 participants). Quality assessment using the CASP/MMAT checklists showed that 80% of studies were of high methodological quality, while 20% were of moderate quality (Table 3). The publication timeline revealed research peaks in 2021 and 2024, suggesting an increasing recognition of AMS as a public health priority in Nigeria. However, a notable gap was identified in 2023, with no eligible studies published during that period, indicating inconsistencies in research productivity.

#### Thematic Analysis: Barriers and Facilitators to AMS Implementation in Nigerian Hospitals

**Theme 1:** Knowledge-Practice Gap. – The Education Crisis in AMS Implementation A recurring theme across the studies was the gap between awareness of AMS principles and their practical application. Deficits in formal education (70%) (Figure 2), inconsistent training, and lack of stewardship curricula perpetuated irrational prescribing and dispensing practices. Even where training programmes existed, they were often short-term, poorly resourced, and insufficient to achieve behavioural change. This reflects a systemic weakness in professional development, necessitating sustained and institutionalised AMS education.

**Figure 2:**
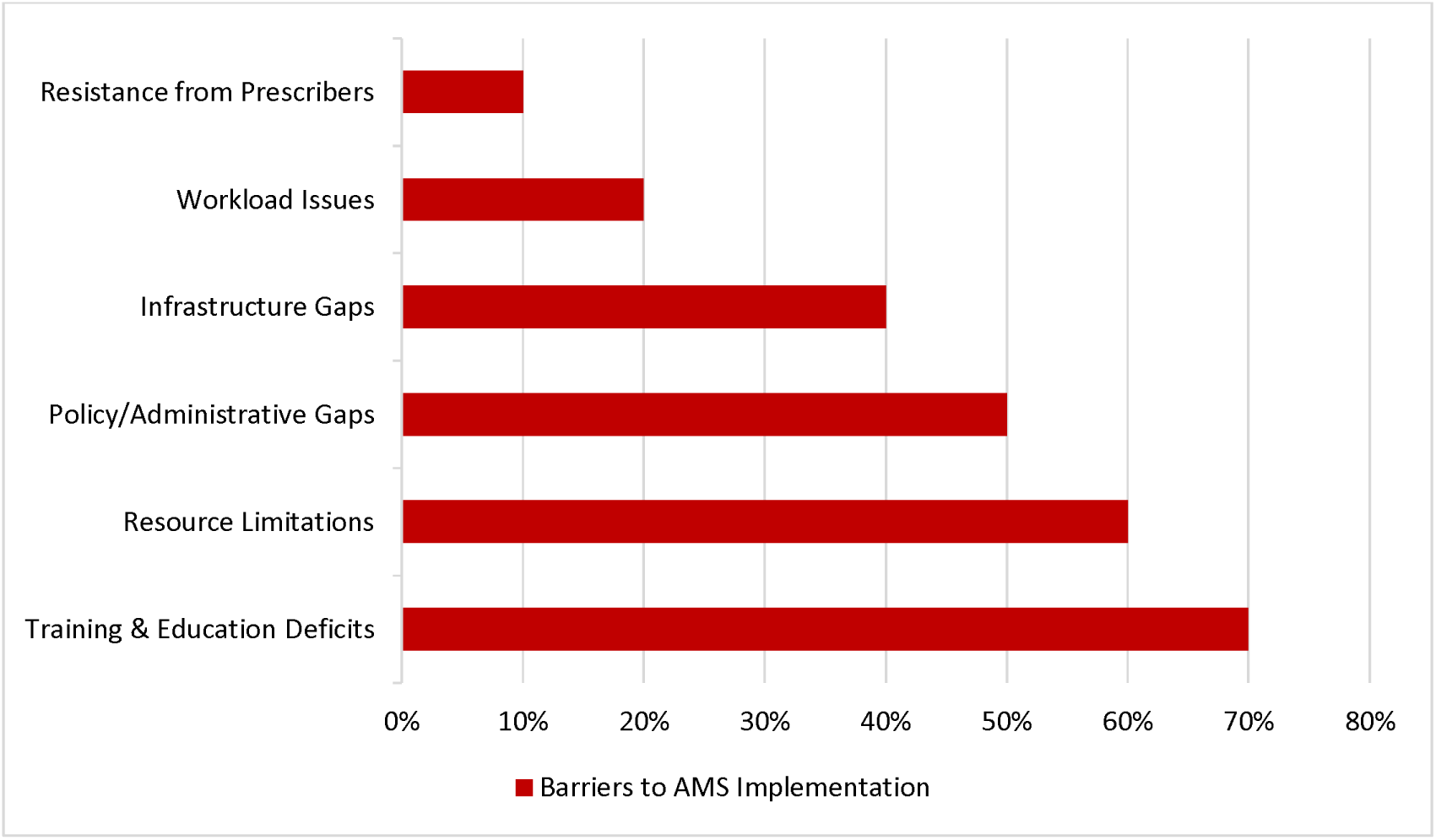
Barriers to AMS Implementation in Nigerian Healthcare Facilities.

**Theme 2:** Organisational Readiness and Leadership – The Foundation for Change in organisational culture and leadership commitment (Figure 3) strongly shaped AMS outcomes. Facilities with leadership support and AMS committees reported more structured and sustained stewardship activities [19]. Conversely, in settings where stewardship lacked administrative backing, interventions were fragmented or donor-driven, limiting their long-term viability. Leadership thus emerges as a linchpin for scaling AMS interventions nationwide.

**Figure 3:**
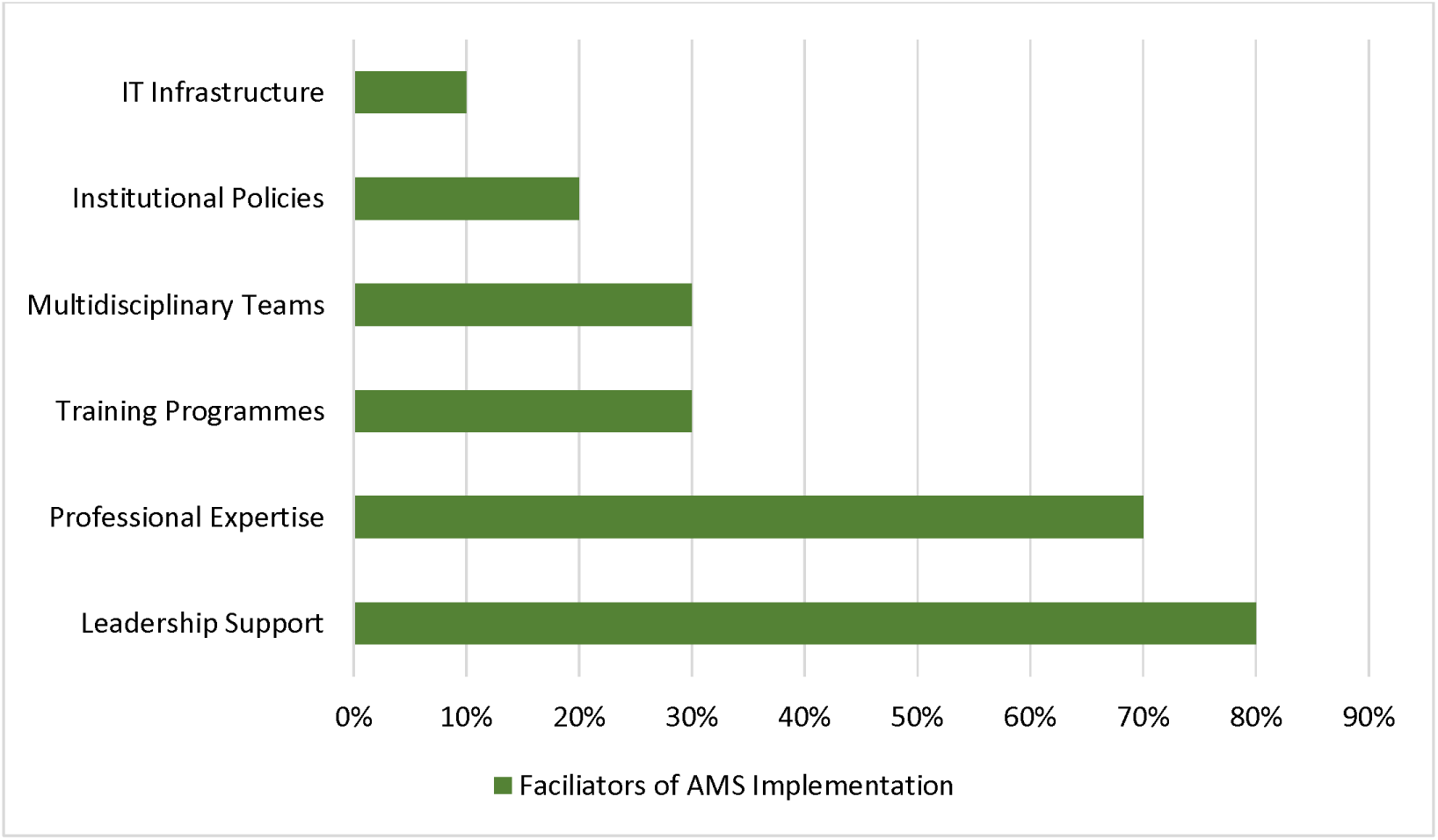
Facilitators of AMS Implementation in Nigerian Healthcare Facilities.

**Theme 3:** Resource Constraints and System Fragmentation - Structural Barriers Multiple studies highlighted weak diagnostic infrastructure, poor funding, and fragmented governance as persistent barriers. These systemic constraints compelled clinicians to rely on empirical prescribing, often with broad-spectrum antimicrobials, reinforcing resistance patterns. The absence of surveillance systems and laboratory integration further weakened stewardship. Without tackling these macro-level deficits, AMS efforts risk being unsustainable and inequitable.

**Theme 4:** The Pharmacist Role in AMS Implementation - Pharmacists emerged as central actors in AMS, particularly in hospital and community pharmacy settings. Their expertise in antimicrobial supply, dosing, and rational use positioned them as key facilitators. However, their role was frequently under-recognised, with limited inclusion in decision-making committees or policy development. Expanding the pharmacist’s role beyond drug supply towards leadership and stewardship functions presents a significant opportunity for strengthening AMS capacity.

#### Integration of Quantitative and Qualitative Findings

The quantitative data highlight the frequency of barriers/facilitators and strategies (Figures 2, 3, & 4), while the qualitative data explains the underlying dynamics. Despite progress in AMS committee formation and education, stewardship is hindered by structural and systemic weaknesses. To achieve a sustainable outcome, a dual focus on strengthening governance and addressing system-level deficits is needed.

**Figure 4:**
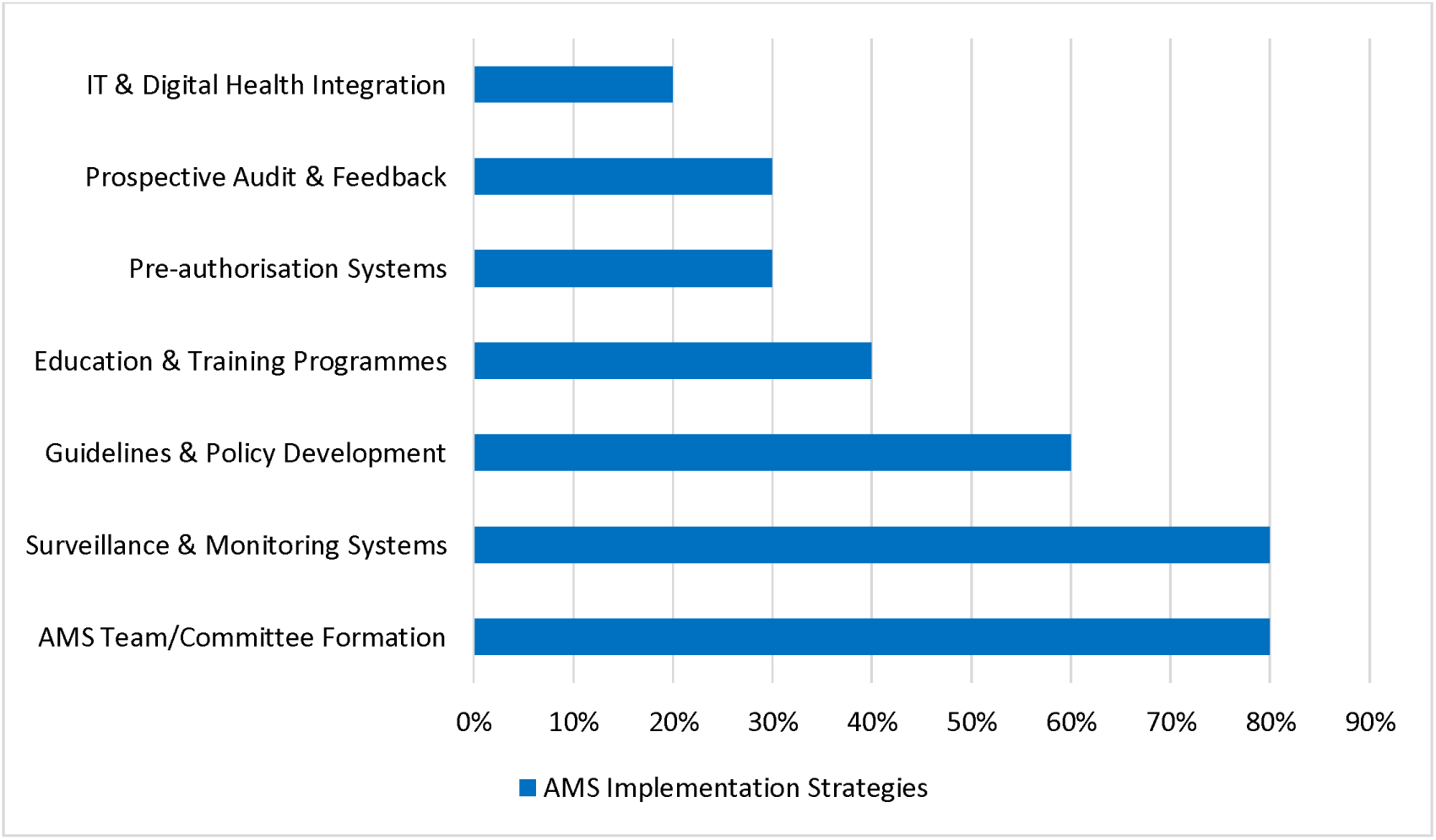
AMS Implementation Strategies Reported in Nigerian Healthcare Facilities.

Figure 5 presents a conceptual framework synthesising the thematic analysis of barriers and facilitators to AMS implementation in Nigerian hospitals. The framework illustrates four interconnected themes that emerged from the qualitative analysis. Theme 1 (Knowledge-Practice Gap) highlights the education crisis, where a 70% deficit in formal education, inconsistent training, and absent stewardship curricula perpetuate irrational prescribing practices, necessitating sustained and institutionalised AMS education. Theme 2 (Organisational Readiness and Leadership) identifies leadership support and administrative backing as critical facilitators, whilst their absence results in fragmented, donor-driven interventions with limited long-term viability. Theme 3 (Resource Constraints and System Fragmentation) underscores structural barriers, including weak diagnostic infrastructure, poor funding, and fragmented governance, which compel clinicians toward empirical prescribing with broad-spectrum antimicrobials, thereby reinforcing resistance patterns. Theme 4 (Pharmacist Role in AMS Implementation) recognises pharmacists as central yet under-recognised actors in stewardship, with significant untapped potential for leadership beyond traditional drug supply functions. The integration node synthesises quantitative findings on barrier frequency with qualitative insights into underlying dynamics, revealing that despite progress in AMS committee formation and educational initiatives, systemic weaknesses persist. The framework converges on a dual-focus solution pathway: strengthening governance structures and resources whilst institutionalising education and expanding pharmacist leadership roles. This integrated approach addresses both immediate implementation challenges and longer-term sustainability requirements for effective AMS programmes in Nigerian healthcare facilities.

**Figure 5:**
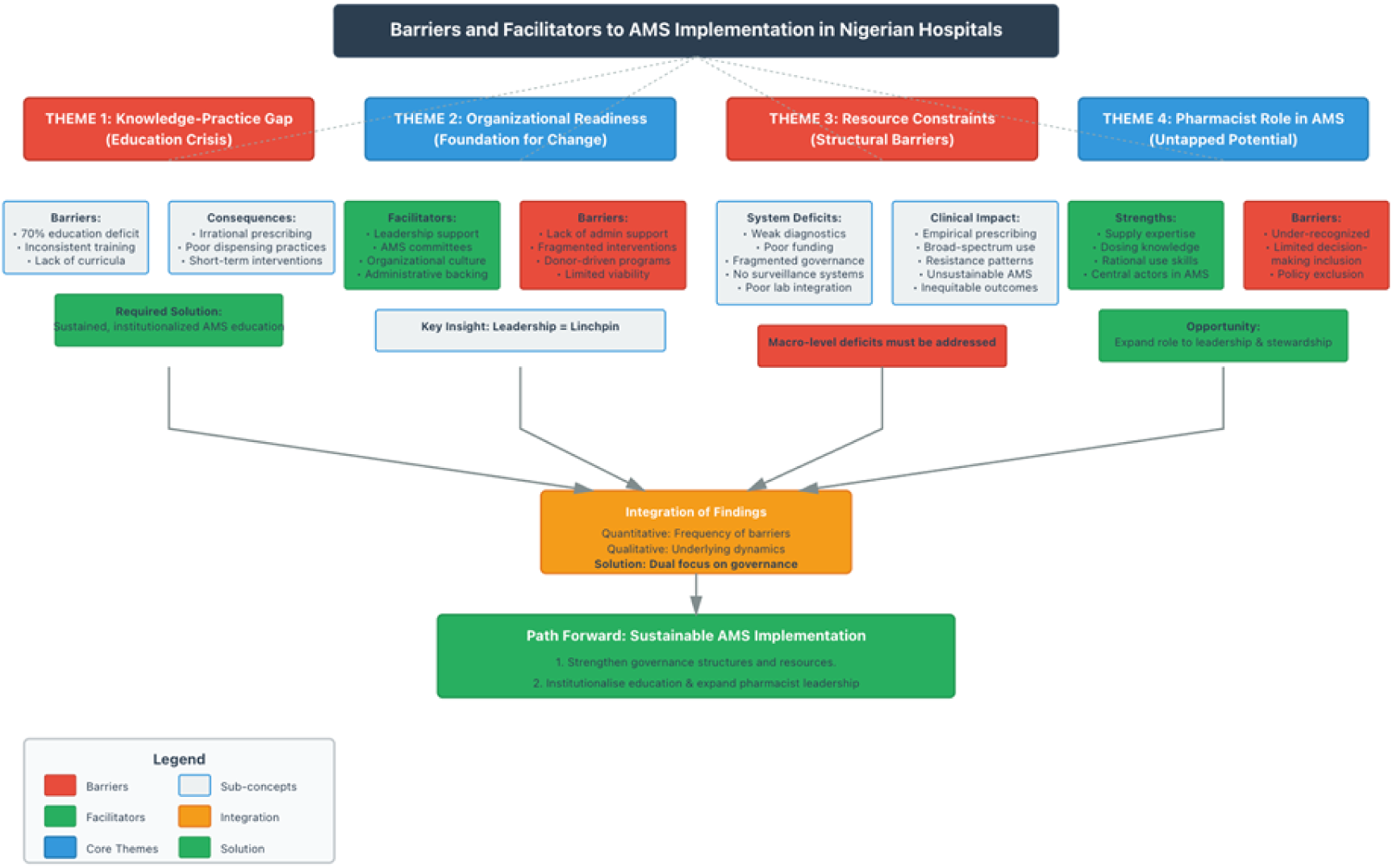
Barriers and Facilitators to AMS implementation in Nigerian Healthcare Facilities.

## Discussion

This systematic review synthesised evidence from ten studies, combining both qualitative and quantitative designs, to explore the barriers, facilitators, and implementation strategies of AMS in Nigerian healthcare facilities. The findings highlight a persistent gap between AMS policy aspirations and real-world practice, underscoring the complex health system challenges that Nigeria faces in addressing AMR. Barriers to AMS Implementation Prescribers and pharmacists frequently identified inadequate laboratory capacity, weak diagnostic stewardship, and insufficient institutional support as key barriers to AMS [18, 19]. These findings align with global research showing that under-resourced laboratory systems and poor diagnostic infrastructure constrain stewardship, particularly in low- and middle-income countries [20, 21]. For example, a systematic review of African AMS programmes found that although some sites achieved improved guideline compliance and reductions in antibiotic use, the overwhelming majority reported limited microbiological capacity, poor data systems and weak policy enforcement [22, 23]. The World Health Organization (WHO) toolkit emphasises a step-wise approach to AMS implementation starting small, building structures, strengthening diagnostics, and engaging champions but warns that resource constraints, poor regulatory environments and absent stewardship committees remain major road-blocks [24, 25]. In Lagos State, Nigeria, Chukwu et al. (2024) conducted a mixed-method survey of prescribers (doctors and pharmacists) to identify facilitators and barriers to AMS implementation. They found that although most respondents acknowledged antibiotic resistance and inappropriate prescribing as problems, major obstacles included inadequate staffing, poor diagnostics, and weak policy enforcement. Key facilitators were stakeholder engagement, established AMS committees, and education/training of prescribers. [26]. Nigeria-specific challenges were also evident, such as erratic medicine supply chains, high patient demand for antibiotics, and profit-driven prescribing practices [27, 28]. These contextual barriers were consistent with the themes identified by Okedo-Alex et al. (2020) [29], who highlighted how systemic weaknesses and irrational antibiotic use undermine AMS effectiveness across sub-Saharan Africa. This corresponds with global evidence: in many LMIC settings, prescribers cite pressured service demands, weak regulatory oversight of antimicrobial distribution, and minimal feedback on prescribing patterns as key impediments to AMS success [30].

### Facilitators of AMS Implementation

Despite persistent barriers, several key facilitators have supported AMS implementation in Nigeria and other LMICs. Strong leadership, multidisciplinary teamwork, and targeted professional training were the most consistent enablers [31]. Hospital administrators showed greater commitment when AMS activities were supported by accountability frameworks and funding [32]. Continuous education, including training on the WHO AWaRe classification, improved prescribers’ stewardship knowledge [18, 27, 33]. Similarly, leadership commitment and AMS committee involvement were major success factors in Nigerian hospitals [26].

Globally, studies confirm that leadership engagement, ongoing training, and local prescribing guidelines enhance antibiotic use and sustainability [23, 34, 35, 36]. Mentorship and “train-the-trainer” models are also effective in resource-limited contexts [24].To maintain progress, AMS must be institutionalised within hospital governance and integrated with infection prevention and control systems [29].

### Challenges of AMS Implementation

Despite notable policy advances, antimicrobial stewardship (AMS) implementation in Nigeria remains hindered by persistent structural, institutional, and behavioural barriers. Core challenges include inadequate diagnostic capacity, limited human resources, weak data and surveillance systems, and erratic antibiotic supply chains [27, 32]. Many hospitals still lack functional microbiology laboratories and electronic systems to produce antibiograms or monitor resistance patterns, perpetuating reliance on empirical prescribing [23, 26].

Although the National Action Plan on AMR 2.0 (2024 - 2028) provides a stronger governance and funding framework, implementation is constrained by weak policy enforcement, limited domestic financing, and poor cross-sector coordination [37, 38]. Financial fragility remains a critical limitation, as most AMS activities depend on inconsistent government allocations or donor support. The absence of a dedicated AMS budget restricts access to laboratory reagents, training resources, and data infrastructure essential for surveillance [38, 39].

Behavioural and cultural factors further undermine stewardship efforts. Patient expectations for antibiotics, profit-driven prescribing in private facilities, and pharmaceutical marketing pressures all contribute to irrational use [28, 29]. Inter-professional hierarchies, particularly between physicians and pharmacists often impede collaboration and feedback mechanisms vital for effective AMS [36]. Governance gaps persist as well; although Nigeria’s first National Action Plan on AMR (2017–2022) prioritised stewardship, enforcement and accountability remain weak due to limited integration into hospital operations and absence of mandatory reporting frameworks [24, 39].

Finally, the COVID-19 pandemic exacerbated these challenges by increasing empirical antibiotic use and diverting diagnostic resources, reversing stewardship gains in many low- and middle-income countries, including Nigeria [38]. These cumulative barriers highlight the urgent need for sustainable financing, stronger governance, and institutional ownership to ensure AMS resilience within the Nigerian health system.

### AMS Implementation Strategies

Nigeria has piloted several antimicrobial stewardship (AMS) implementation strategies with varying degrees of success. Commonly adopted approaches include prescriber education, audit and feedback mechanisms, antimicrobial prescribing guidelines, and point prevalence surveys [28, 29, 40, 41]. In this review, the three most frequent strategies were AMS team or committee formation (80%), education and training programmes (70%), and development of prescribing guidelines and hospital policies (60%). These align closely with the National Action Plan on Antimicrobial Resistance (2017-2022), which identified governance, workforce training, and surveillance as the core pillars of AMS implementation [39]. To build on these foundations, Nigeria launched the National Action Plan on Antimicrobial Resistance (NAP 2.0; 2024 - 2028) in October 2024, adopting a One Health framework that integrates human, animal, and environmental health sectors [37]. NAP 2.0 introduces a costed implementation roadmap, a national monitoring and evaluation system, and enhanced coordination across ministries. These reforms mark a strategic transition from fragmented, donor-driven projects to a more institutionalised governance model aimed at achieving long-term sustainability and national ownership.

At the operational level, hospitals with established AMS committees demonstrated improved antibiotic review processes, regular prescriber feedback, and integration of stewardship indicators into facility performance monitoring [26]. Educational interventions, such as continuing medical education, in-service pharmacy training, and stewardship workshops, significantly enhanced prescribers’ confidence and adherence to rational antibiotic use [28, 29]. The adoption of local antibiotic formularies and adaptation of the WHO AWaRe classification further promoted treatment standardisation and optimised antibiotic selection in tertiary care settings [24].

Nevertheless, sustainability remains a persistent challenge. Many AMS programmes in Nigeria continue to rely heavily on donor funding or external technical assistance [23]. This dependence raises concerns about scalability and programme continuity once external support ends. Weak domestic financing structures and the absence of dedicated AMS budget lines within hospitals further constrain local ownership and accountability [34]. Strengthening domestic funding, embedding AMS priorities within national health financing frameworks, and integrating stewardship into broader quality improvement and infection prevention systems are therefore critical to ensuring the long-term success of AMS initiatives in Nigeria.

### Thematic Insights

The thematic synthesis of the included studies revealed four interconnected themes shaping antimicrobial stewardship (AMS) implementation in Nigeria. A persistent knowledge–practice gap was evident: although awareness of stewardship principles and the WHO AWaRe classification has grown, translation into consistent clinical practice remains limited. This disconnect reflects inadequate ongoing training and resistance to behavioural change among prescribers, echoing findings by Okedo-Alex et al. (2020), who highlighted the urgent need for structured stewardship education and institutional learning frameworks. Comparable studies from other low- and middle-income countries (LMICs), including Kenya and India, have similarly noted that knowledge improvements alone do not guarantee appropriate antibiotic use without systems that reinforce accountability and peer feedback [23, 32].

Organisational readiness and leadership engagement emerged as another critical determinant of AMS success [17]. Facilities with strong administrative support, clear accountability mechanisms, and multidisciplinary governance structures demonstrated greater progress in embedding stewardship into routine clinical workflows [26]. This pattern aligns with international evidence showing that visible leadership commitment and institutional governance are foundational for cultural change and long-term stewardship sustainability [24].

Resource constraints and system fragmentation remained recurring barriers, with most facilities reporting limited diagnostic capacity, unreliable laboratory networks, and erratic antibiotic supply chains [28, 42]. Such structural weaknesses mirror global findings that inadequate laboratory infrastructure and supply instability undermine data-driven decision-making and surveillance, particularly in LMICs.

Pharmacists consistently emerged as key AMS champions, often leading prescription reviews, audit feedback, and training initiatives [28, 29, 40]. This reinforces the global trend recognising pharmacists as pivotal in multidisciplinary stewardship teams, providing clinical oversight and bridging communication gaps between prescribers and policy implementers [24]. Collectively, these thematic insights underscore that sustainable AMS implementation in Nigeria, and across similar resource-constrained settings, depends on integrating knowledge with leadership, infrastructure, and inter-professional collaboration [34].

The findings have important implications for strengthening Nigeria’s health system. Although the country demonstrates strong professional capacity and commitment to implementing antimicrobial stewardship (AMS), urgent systemic reforms are needed to address weaknesses in diagnostics, fragmented governance structures, and inconsistent funding. To achieve sustainable progress, this review highlights several key recommendations. First, policy reform is essential to establish stronger national and state-level AMS policies supported by clear accountability frameworks. Second, capacity building should focus on expanding prescriber and pharmacist training, including integrating AMS principles into preservice education. Third, adequate resource allocation is required, with investment in diagnostics, laboratory systems, and digital health infrastructure to strengthen AMS implementation. Fourth, AMS efforts should be closely integrated with infection prevention and control (IPC) programmes to enhance coordination and maximise health system efficiency. Finally, long-term sustainability planning is vital, with an emphasis on reducing donor dependency through government-led financing and stronger domestic ownership of AMS initiatives.

Future research should prioritise multi-centre studies that include rural and private healthcare facilities, which remain underrepresented despite accounting for a significant proportion of antibiotic use. There is also a need for long-term monitoring and cost-effectiveness analysis to inform policy and financing decisions. Importantly, integrating stewardship into wider infection prevention and control (IPC) programmes and embedding AMS principles into preservice curricula for health professionals will be critical for sustainability in Nigeria.

### Strengths and Limitations

This review has several strengths. First, it is the first systematic review to synthesise evidence on AMS implementation in Nigeria, providing timely insights into an underexplored area. Second, the inclusion of diverse study designs (quantitative and qualitative) allowed for triangulation of findings and improved depth of analysis. Third, the review incorporated evidence from both hospitals and community pharmacies, broadening its relevance beyond tertiary facilities alone.

Nevertheless, limitations must be acknowledged. The evidence base remains limited, with only ten studies meeting inclusion criteria, which restricts the breadth of available insights. Second, the majority of included studies were based in tertiary and urban hospitals, limiting generalisability to primary care and rural contexts, where much antibiotic use occurs. Finally, potential publication bias is likely, as unsuccessful or less visible AMS initiatives may remain unpublished, potentially skewing the findings towards positive outcomes.

### Conclusion

This systematic review highlights the barriers, facilitators, and strategies influencing AMS implementation in Nigeria. The main barriers identified were inadequate training and education, limited diagnostics, and weak institutional policies. The main facilitators were leadership support, professional expertise, and multidisciplinary collaboration. The most common implementation strategies included AMS committee formation, prescriber education programmes, and prescribing guidelines.

From the thematic synthesis, the knowledge–practice gap emerged as the dominant theme, reflecting challenges in translating awareness into consistent AMS practice. Despite strong human resource potential, Nigeria faces structural barriers that limit sustainability. Embedding AMS into wider health system reforms, including governance, infrastructure, and financing, is essential for long-term success.

Future research should target rural and private-sector facilities, evaluate cost-effectiveness, and explore integration with IPC programmes, ensuring AMS is institutionalised through policy enforcement, stable financing, and professional education.

### Availability of Data

All data supporting the conclusions of this article are included within the article and a list of references.

## Abbreviations

AMR: Antimicrobial Resistance
AMS: Antimicrobial Stewardship
ASP: Antimicrobial Stewardship Programme
AWaRe: Access, Watch and Reserve
CASP: Critical Appraisal Skilled Programme
CI: Confidence Interval
CPD: Continuing Professional Development
DDD: Defined Daily Dose
DDI: Drug-Drug Interaction
DOT: Days of Therapy
DTC: Drug and Therapeutics Committee
ID: Infectious Diseases
IPC: Infection Prevention and Control programme
IT: Information Technology
M&E: Monitoring and Evaluation
MMAT: Mixed Method Appraisal Tool
N/A: Not Applicable
N: Number
NAFDAC: National Agency for Food, Drug and Control
NCDC: Nigeria Centre for Disease Control
OR: Odd Ratio
PCN: Pharmacists Council of Nigeria
PICO: Population, Intervention, Comparison, and Outcome
PPS: Point Prevalent Survey
PRISMA: Preferred Reporting Items for Systematic Reviews and Meta-Analyses
UH: University of Hertfordshire
UK: United Kingdom
UN: United Nations
WHO: World Health Organisation

## Acknowledgements

The authors would like to acknowledge the Academic Support team from the University of Hertfordshire (UH), who helped in reviewing this study.

## Author Information

### Ethics Declaration

Not applicable.

### Consent for publication

Not applicable.

### Data Availability Statement

All data produced in the present study are available upon reasonable request to the authors

